# Facilitators and Barriers to Patient-Caregiver Dyadic Recruitment in Transplantation

**DOI:** 10.1101/2025.01.13.25320484

**Authors:** Brittany Koons, Rachel Lehman, Barbara Riegel, Harleah Buck

**Affiliations:** M. Louise Fitzpatrick College of Nursing, Villanova University, Villanova, PA, USA; University of Pennsylvania School of Nursing, Philadelphia, PA, USA; Senior Research Scientist, Center for Home Care Policy & Research at VNS Health, New York, NY; College of Nursing, The University of Iowa, Iowa City, Iowa, USA

## Abstract

**Background:** Despite growing awareness of the dyadic role in transplant care and a mandate for patient–caregiver dyads for transplant listing, the integration of dyadic science into transplantation research is lacking. Recruiting transplant patient-caregiver dyads has unique challenges that need to be considered when designing and conducting dyadic studies in transplantation.

**Objectives:** To present 1) the barriers and facilitators to dyadic recruitment in a patient-caregiver transplant population that we encountered and 2) strategies developed to overcome these challenges.

**Methods:** We used the Social Marketing Mix framework to guide this methodological report of patient-care dyadic recruitment strategies employed during a post-lung transplant psychometric study of 50 patient-caregiver dyads.

**Results:** We identified several facilitators of dyadic recruitment in the lung transplant population including: 1) conducting a study of high relevance to both patients and caregivers, which helped to facilitate maximum engagement of participants, 2) using remote recruitment and data collection strategies to improve accessibility to participation and to minimize the amount of time or energy required to participate, 3) conducting patient and caregiver study visits independently from one another, which allowed for scheduling flexibility, and helped improve participation among dyad members who do not live together, and 4) establishing clinical partnerships and having acquired clinical experience with the target population. We also identified barriers to dyadic recruitment that require careful planning in future studies including: 1) high health care utilization, which can delay the recruitment timeline, 2) recruiting patients and caregivers independently within relevant timeframes, 3) gatekeeping, when the patient or the caregiver block researcher access to the other dyad member, 4) establishing contact with the participant via the phone, and 5) limited study staffing that reduced recruitment and study visit scheduling flexibility.

**Discussion:** To our knowledge, this is the first methodological report to present the barriers and facilitators to dyadic recruitment in a patient-caregiver transplant population. Our experience and lessons learned can be used to inform future research teams to successfully design and conduct much needed dyadic research in organ transplantation.

## 1. Introduction

Organ transplantation prolongs life expectancy and improves quality of life for patients with end-stage organ failure.^1^ More than 150,000 solid organ transplants are performed annually worldwide.^2^ However, more than 10% of patients with organ failure who need a transplant are not eligible to be listed for one because they do not have adequate caregiver support, a long-standing absolute contraindication to transplant listing.^3^ The caregiver criterion is predicated on an assumption that robust caregiver support promotes improved transplant outcomes by providing patients with support that is critical for adherence to the complex post-transplant self-care regimen.

Self-care involves three key behaviors— maintenance, monitoring, and management— that aim to maintain physiologic and mental stability, facilitate symptom monitoring, and optimize chronic illness management.^4^ Patient self-care is critical to transplantation success but challenging. For example, after lung transplant, patients are expected to take numerous prescribed medications (i.e., >40 medications a day) and attend many clinic appointments (self-care maintenance), monitor lung function daily (self-care monitoring), and immediately communicate clinical changes to their transplant team (self-care management).^5^ Due to the high burden of transplant self-care and because poor self-care is a risk factor for post-transplant rejection, poor health related quality of life, and death,^6,7–9^ verification of a primary caregiver is an absolute requirement for transplant listing.^10,11^ Yet, transplant research has mostly focused on patient contributions to their own self-care and has rarely considered caregiver contributions to transplant self-care. Thus, the evidence base to support the caregiver requirement for transplant listing is weak.^12 13–15^ Further, despite this mandate for patient-caregiver dyads in transplantation, we know very little about how patient-caregiver dyads engage in and manage transplant self-care together and how dyadic engagement in transplant care affects outcomes.^13^

Given the critical importance of patient-caregiver dyads in transplantation and the limited integration of dyadic science in this field, studies of patient-caregiver dyads have been identified as a priority for transplantation research.^13^ However, recruiting transplant patient-caregiver dyads has unique challenges to consider when designing and conducting dyadic studies. To our knowledge, there is no literature describing patient-caregiver dyadic recruitment methods in the transplant population. The purpose of this paper is to provide a methodological report of dyadic recruitment strategies used during a post-lung transplant psychometric study. Using the Social Marketing Mix framework^16^ we will describe: 1) the barriers and facilitators to dyadic recruitment that we encountered, and 2) strategies we developed to overcome these challenges.

### Overview of the parent study

We conducted a cross-sectional study of post-lung transplant patient-caregiver dyads recruited from a large academic transplant center located in an urban city in the United States. The goal of this study was to psychometrically evaluate a dyadic measure that evaluates how patients and caregivers engage in patients’ self-care. This dyadic measure, formerly called the Dyadic Symptom Management Type scale, was originally developed and validated for use in in heart failure population.^58^ With permission from the original author, we revised this tool for use in lung transplantation, renamed it the Dyadic Self-Care in Transplant (D-SCIT) scale, and tested its psychometric properties. We assessed face validity using cognitive interviewing and convergent and divergent validity by testing the relationship between the D-SCIT and mutuality, a closely related concept measuring dyadic relationship quality. We recruited post-lung transplant patient-caregiver dyads who were adults (age > 18 years), spoke English, and were >6 months from lung transplant surgery. This study was approved by the Institutional Review Board (IRB) of the university (Protocol # 833262).

#### Original Recruitment Methods and Data Collection

The principal investigator (PI) screened the electronic medical record weekly for lung transplant recipients who met eligibility criteria. The PI and a trained research assistant (RA) then had a weekly phone call to discuss the list of patients to contact. Then, the RA called eligible patient participants to discuss the project and solicit participation. A phone script was used to standardize the recruitment approach. Patients who were willing to participate and who could identify a caregiver support person to participate then provided verbal informed consent. The RA then administered the study instruments (i.e., D-SCIT and Mutuality Scale) and conducted the cognitive interviewing with the patient. Then, the RA contacted the caregiver and followed the same procedures.

#### Recruitment and Enrollment Results

We enrolled 60 patients and 50 caregivers in the study— a total of 50 complete patient-caregiver dyads. Our study flow diagram (**Figure 1**) presents data on participants who were recruited and enrolled. Overall, we enrolled 75% of the patients who we established contact with. We attempted to recruit 60 caregivers of patients enrolled in our study and ultimately, we were able to enroll 84% of these caregivers.

**Figure 1.**
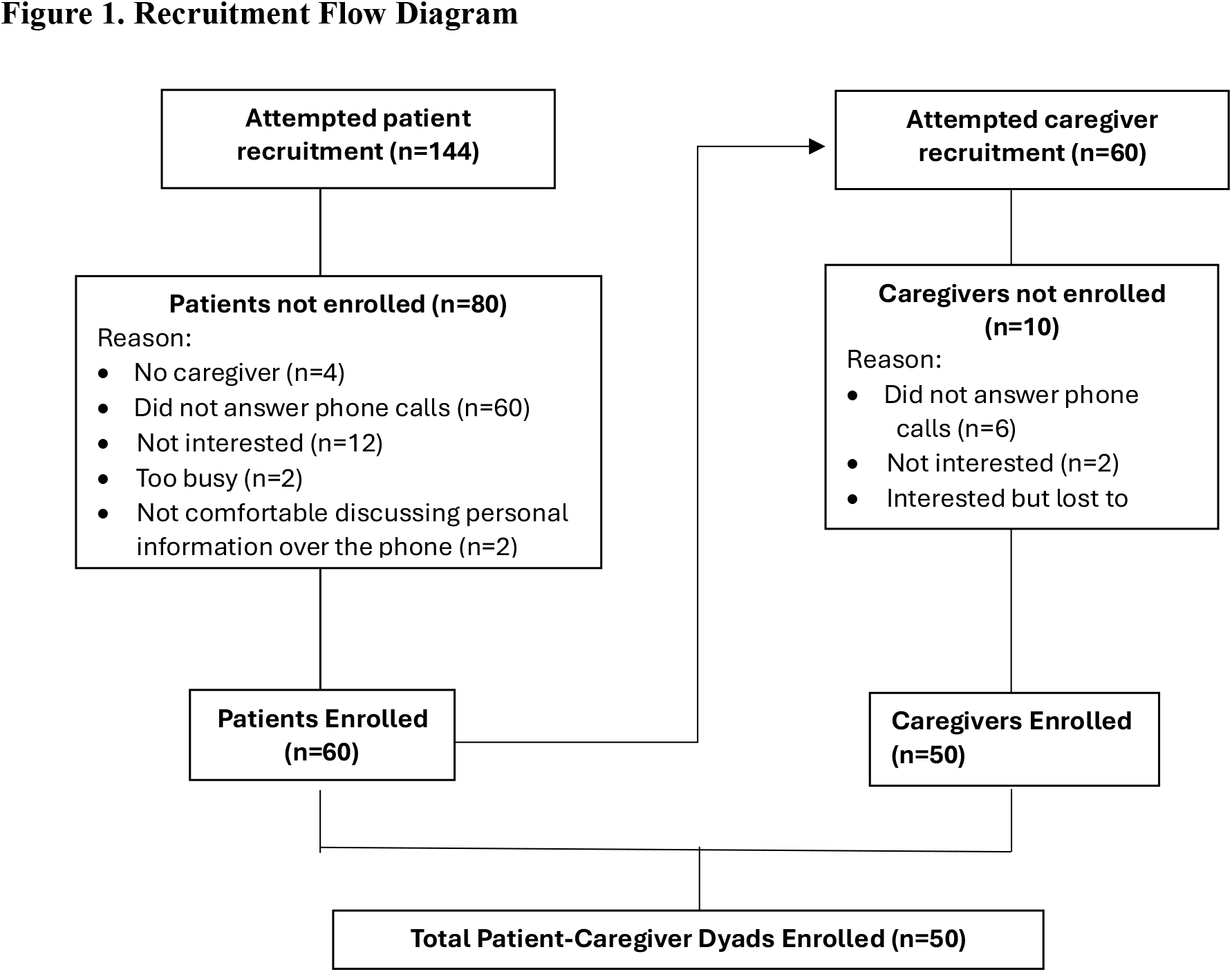
Recruitment Flow Diagram.

## 2. Methods

We used the Social Marketing Mix (SMM) framework^16^ to guide this methodological report (see Table 1 for domains and definitions). The SMM framework highlights important concepts for researchers to consider when planning and implementing recruitment strategies for clinical research studies. This framework has been effectively used to report successful patient recruitment strategies and challenges encountered in various clinical settings, such as palliative care and hospice,^17–20^ but has not yet been used to report dyadic recruitment strategies in the transplant population.

**Table 1.**
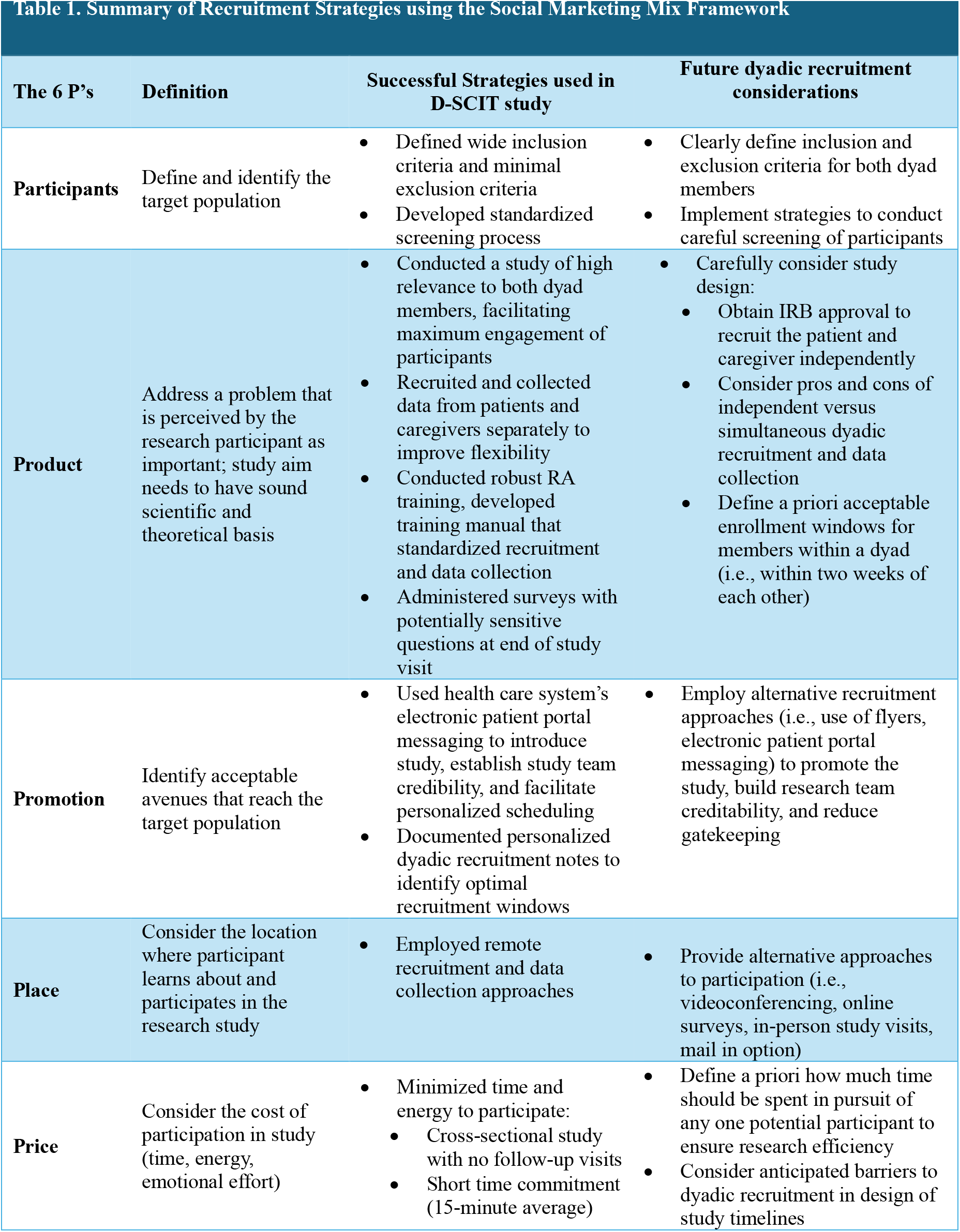
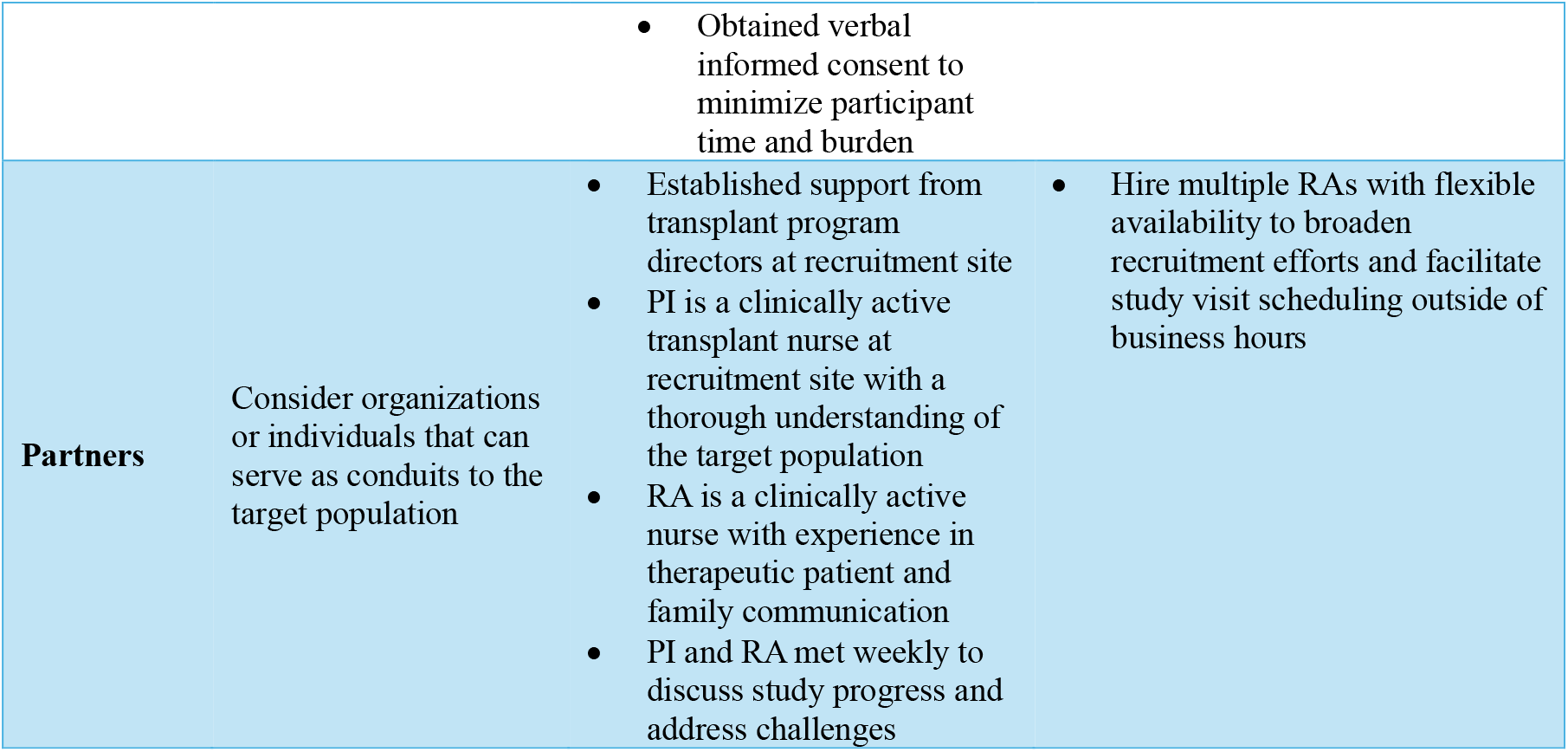
Summary of Recruitment Strategies using the Social Marketing Mix Framework.

### Results

The results are organized to address our two aims: to report 1) barriers and facilitators to dyadic recruitment that we encountered, and 2) strategies we developed to overcome these challenges according to the SMM framework.

### Participants: defining the target population

#### Barrier: High health care utilization

The lung transplant population has high health care utilization, especially in the first postoperative year, including unplanned hospitalizations, scheduled clinic visits, and planned procedures (i.e. bronchoscopy). Some participants (n=4) were ineligible to participate because they were hospitalized and therefore, we were either unable to contact the patient, or the patient was not well enough to participate. Several patients (n=8) had scheduled clinic appointments causing patients to be temporarily unavailable and our team to delay recruitment calls.

#### Facilitator: Inclusivity

We clearly defined study eligibility for patients and caregivers prior to the start of the study. We had wide inclusion criteria and minimal exclusion criteria. Broad inclusion criteria helped to facilitate patient recruitment by ensuring a high percentage of patients who were screened were eligible to participate in the study.

#### Strategy: Standardized screening procedures

The PI reviewed the electronic health record to ensure that phone calls were not made to patients around planned admissions (i.e., planned bronchoscopy), scheduled clinic visits, or to patients who were actively hospitalized, recognizing that these were not opportune times for patients to engage in research and investment of study resources to recruit patients during these times would likely not be successful. One of our inclusion criteria was that both the patient and their primary caregiver had to be willing to participate. Prior to enrollment, we asked the patient if they had a caregiver who we could contact. Despite the requirement that patients identify a primary caregiver before transplant, four patients could not identify a primary caregiver after transplant and were therefore, not enrolled in the study. Our screening procedures proactively protected the patient from unnecessarily investing their time into participating in a study that they were ultimately not eligible to participate in because they could not identify a caregiver and prevented our study team from investing time and resources into recruiting ineligible or unavailable transplant patients.

### Product: study aims and design

#### Barrier: Timing

We designed our study to recruit and enroll patients and caregivers separately. Although this approach has advantages, it also has distinct challenges. After enrolling the patient, we found it difficult to recruit and enroll several patients’ caregivers within a timeframe relevant to the study. For example, among four dyads, significant time had passed since the patient’s study visit before the caregiver was enrolled in our study (> 30 days). There were 10 caregivers who we were unable to recruit and enroll into the study despite concerted efforts by the RA; this required considerable investment of study time and resources with ultimately, no success.

#### Facilitator: Relevancy of study

Only a small number of patients (n=12) and caregivers (n=2) declined to participate in the study because they were not interested. We found that the aim of our study was of high relevance to both patients and caregivers. This helped to facilitate maximum engagement of participants.

#### Facilitator: Participant enthusiasm

We found that lung transplant patients were accustomed to and receptive to study recruitment calls and often, eager to engage in lung transplantation research. They frequently reported thankfulness for the care they received during their transplant journey and expressed a sense of wanting to “give back.”

#### Facilitator: Flexibility

Recruiting, enrolling, and collecting data from dyad members separately was a study design feature that we found resulted in more benefits than barriers. This design allowed for flexible recruitment and study visit scheduling that centered around the participant’s availability. This design also allowed participation among dyad members who do not live together; 16% of dyads enrolled in our study were either parent-child (n=7) or patient-sibling (n=1) dyads who did not live together.

#### Strategy: Training

A training manual was developed that described inclusion and exclusion criteria, accrual goals, screening and recruitment procedures, and provided standardized scripts for recruitment. This training manual was used by the PI to train the RA and served as a helpful resource for the RA throughout the study. Additional RA training included how to address participants’ questions about the purpose of the study and how to explain the meaning of the survey questions and response options. The RA was well prepared to clarify participants’ questions or concerns related to the study purpose and survey questions. For example, one of the instruments, the Mutuality Scale, asks the participants questions about their relationship with the other dyad member. Some participants questioned the relevancy of this measure, and the RA was well prepared to clarify participants’ questions or concerns related to the meaning and purpose of the survey items. Additionally, we intentionally administered this survey last to allow the RA to develop rapport with the participant prior to administering this survey that asks questions that some participants may perceive as personal or sensitive.

### Promotion: recruitment approaches

#### Barrier: Gatekeeping

Gatekeeping occurs when the patient or the caregiver block researcher access to the other dyad member. A few patients felt that their caregiver was either too busy or would not want to participate in research and asked us not to contact their caregiver. Likewise, a few caregivers answered the patient’s phone and blocked researcher access to the patient, often citing that the patient gets too many phone calls or that they felt that the patient would not be interested in participating in research. Additionally, the target for recruitment for our IRB-approved protocol was the patient and we relied on the patient to provide the name and contact information for their primary caregiver. An unanticipated consequence of this decision is another form of gatekeeping; our protocol may have limited our recruitment to patients who were independent and motivated to engage in research and restricted our ability to recruit eligible caregivers of sicker patients who may have been interested in participating.

#### Strategy: Strategic recruitment planning

To optimize our reach to the target population, our study team kept detailed, personalized recruitment notes for each patient and caregiver in REDCap, a secure data management system. Notes included when calls were made, the best time to call patients and caregivers, and what days or times to avoid calls; this helped our team to strategize recruitment calls. We also developed a standardized script that was used by the RA for all study contacts to ensure delivery of understandable and consistent messaging.

### Place: accessibility to participation

#### Barrier: Remote recruitment and data collection

Our greatest barrier to recruitment was establishing remote contact with the patient and caregiver. Many patients and caregivers were difficult to reach by phone and calls often went to voicemail. Ultimately, we were unsuccessful in recruiting 60 eligible patients and 6 caregivers of patients who were enrolled in the study because we could not establish contact by phone. We learned that patients and caregivers often screen their phone calls and do not answer calls from unfamiliar phone numbers. When cold calling potential participants, we found it more challenging to build creditability and establish trust. As a result, two patients declined participation because they did not feel comfortable sharing information over the phone.

#### Facilitator: Remote recruitment and data collection

Although we encountered barriers to our remote recruitment and data collection approaches, there were also advantages that greatly facilitated dyadic recruitment and enrollment in our study. All study activities were conducted over the phone at a time convenient for the participant rather than requiring in-person participation, access to a computer, or placing the burden on the participant to obtain, complete, and send back mail-in surveys. Additionally, our remote recruitment and data collection approach made participation more accessible and less burdensome for many patients and caregivers who are older or who do not live close to the transplant center.

#### Strategy: Employing alternative recruitment approaches

We began to use the health care system’s secure online patient portal to message patients, informing them about the purpose of the study, and that they may receive a phone call from the RA (providing the RA’s name and area code). If willing to participate, we encouraged the patient and caregiver to reply to our message indicating the best day and time that we could contact them. Implementing this recruitment approach helped to raise awareness about the study, establish the research team’s credibility with potential participants, provided participants the opportunity to express willingness to participate in the study, and facilitated personalized study visit scheduling.

### Price: cost of participation

#### Barrier: Time commitment

Although the time commitment to participate in this study was minimal, for some patients and caregivers, this was a barrier to participation. We also had four patients and two caregivers who expressed interest in participating but were lost to follow-up to schedule a remote study visit.

#### Facilitator: Minimizing participant time and energy

Our study required no follow-up study visits. All study visits were conducted over the phone, at a time convenient for the participant, and no travel was required. Our approach to informed consent minimized participants’ time and burden. We reviewed the consent form with the participant over the phone and obtained verbal informed consent from the participant. This approach eliminated the extra steps required to obtain written consent, allowed for quick progression to study questions, and decreased total call time. Our study visits were relatively short; patient study visits averaged 16 minutes (range, 10-45 minutes) and caregiver study visits averaged 15 minutes (range, 7-45 minutes). These aspects of our study design minimized the amount of time and energy (costs) required to participate, therefore, facilitating dyadic recruitment and enrollment.

### Partners

#### Barrier: Study staffing

Adequate study staffing is critical for study success. For this study, only one RA conducted the study visits, and we were limited in our ability to make calls to participants in the evenings or over the weekend. Study calls were made during business hours, when patients often had other appointments or when some patients and caregivers were working. Limiting calls to business hours may have slowed the recruitment process or prohibited us from recruiting and enrolling patients and caregivers who were unable to answer their phone or participate during the weekday.

#### Facilitators: Clinical Partnerships and clinical experience

We developed a partnership with the recruitment site. The PI introduced the study objectives to transplant team directors at the hospital and obtained approval to conduct the study. The RA and PI have experience working with chronically ill patients and families; the RA is a clinically active nurse with over a decade of experience in therapeutic patient and family communication. The PI is a clinically active nurse who cares for post-transplant patients at the hospital. Our clinical partnership and team’s experiences with building relationships with patients and caregivers helped to inform the study design and facilitated implementation of the research study.

#### Strategy: Study team meetings

To improve efficiency in recruitment, enrollment, and data collection, the PI and RA met weekly to discuss study progress and address challenges in real time using collaborative problem-solving.

## 3. Discussion

Patient-caregiver dyadic studies in lung transplantation are rare but needed given the requirement for transplant eligibility and the critical role of the dyad in managing life-long post-transplant care. The purpose of this paper is to provide a methodological report of post-lung transplant patient-caregiver dyadic recruitment. In our study, we achieved higher recruitment rates (75% of patient contacts and 84% of caregiver contacts) than those reported for dyadic studies in other clinical specialties such as hospice (32%-42%),^18,21^ heart failure (37%-59%),^22,23^ cancer (68%),^24^ and dementia (27%)^25^ suggesting that the recruitment strategies we have developed are powerful tools for use in dyads. That said, we learned additional recruitment strategies along the way which we would like to share with others interested in conducting dyadic research. Here, we provide an overview of “lessons learned” from our dyadic recruitment and potential strategies to overcome barriers to dyadic recruitment in future research (**Table 1**).

### Lessons Learned and Strategies for Overcoming Barriers to Dyadic Recruitment Lesson one

We found that recruiting and enrolling a dyad is more complex than enrolling an individual. Our study aimed to recruit and enroll four types of dyads characterized by how patients and caregivers engage in the patient’s transplant self-care—collaborative, with both members engaging in transplant self-care, patient-or caregiver-oriented, with either the patient or caregiver solely engaging in transplant self-care, or incongruent with dyads disagreeing on who engages in transplant self-care. We enrolled only two dyads that were caregiver-oriented, meaning the caregiver took care of most of the patient’s transplant care with little input from the patient. Our approach of first contacting patients before recruiting caregivers may explain why we were unable to recruit a higher number of caregiver-oriented dyads, who may have been more likely to answer the phone than the patient they cared for. *Suggested strategies:* depending on the aims of the study, researchers should consider obtaining IRB approval to recruit the patient or caregiver, without relying on one dyad member to provide access to the other dyad member. Also, researchers need to implement strategies to conduct careful screening of participants to avoid recruiting patients who are either not eligible to participate because they do not have a transplant caregiver who can participate and to avoid recruiting patients and caregivers during times that may be perceived as stressful (i.e., hospitalizations, scheduled procedures); this is especially important when recruiting from a chronically ill patient population. Researchers also need to consider the health care utilization rates in the population they are recruiting from because high health care utilization rates, as seen in the transplant population, can slow down the recruitment process and this factor needs to be considered when developing study timelines.

### Lesson two

Our greatest barrier to recruitment was establishing contact, via the phone, with the patient and caregiver. We also found that some patients and caregivers were uncomfortable sharing information over the phone. These findings highlight the need to identify additional ways to promote the study and alternative approaches to administering surveys. *Suggested strategies:* our approach in using the health care system’s online patient portal was highly successful in recruiting patient and caregivers and facilitating convenient scheduling. Using the secure online portal helped our team build credibility and established trust with the participant. Developing a flyer for distribution in the clinic to introduce the study purpose and the research team may be an additional approach to promote the study and build creditability with potential participants. These alternative approaches to promoting the study may also help to alleviate gatekeeping by exposing potential patient or caregiver participants to the study who otherwise may not have been presented with the opportunity to participate. Offering alternative approaches to administering surveys either via videoconferencing (e.g., Zoom), secure online data capturing systems (i.e., REDcap), in-person study visits, or a mail in option, may help to recruit those participants who have concerns sharing information over the phone.

### Lesson three

Our study’s design to recruit and conduct study visits with patients and caregivers separately allowed for more flexibility in recruitment, enrollment, and data collection. However, this approach also raised some challenges. For some dyads, the number of days between study visits for members within the dyad was prolonged. The time difference between patient and caregiver study visits did not impact the aims of our study but may be important to consider during the design phase in future dyadic studies. *Suggested strategy:* researchers need to consider the aims of their study and whether time has a significant impact the outcomes of interest. This may be particularly important for longitudinal studies with repeated measures. This issue needs to be carefully considered in the study design and acceptable enrollment windows for members within a dyad should be defined prior to the start of the study. Additionally, in some cases, we were unable to recruit the patient’s caregiver either because the caregiver did not answer their phone despite concerted efforts by the RA or the caregiver expressed interest but did not follow through with scheduling a study visit. These issues raise an important question about the acceptable costs to conduct the study. Namely, how much time and resources the research team should invest in recruiting participants, especially the other dyad member, something that we had not defined at the start of our study. When designing studies and training staff for data collection, it is important to define how much time should be spent in pursuit of any one potential participant to ensure research efficiency. If this becomes a persistent challenge, some studies have had better success restricting dyadic recruitment (i.e., recruiting both dyad members simultaneously) which may result in less missing data, but at the expense of less flexibility in enrollment and data collection.^23^ Lastly, researchers need to consider the increased time investment required to recruit dyads, when planning study timelines to ensure study feasibility.

### Lesson four

Our study was constrained by only having one RA to recruit, enroll, and collect data. This may have limited our ability to maximize flexibility for recruitment and scheduling study visits. *Suggested strategy;* plan to hire multiple trained RAs to undertake recruitment efforts, including during evening and weekend hours; this could greatly increase recruitment rates and decrease missing data. The need to hire multiple RAs to successfully recruit and enroll dyads requires funding, an important consideration for grant budgets.

## Conclusion

Our hope is that our experience and lessons learned will prepare future research teams to successfully design and conduct much needed dyadic research in organ transplantation. While dyadic research can be more challenging than conducting individual patient-or caregiver-focused studies, the dyadic perspective can meaningfully shape clinical transplant care and outcomes for thousands of patients and caregivers who are waiting for transplant and responsible for life-long post-transplant care.

## Data Availability

All data produced in the present work are contained in the manuscript

## Notes

### Competing Interest Statement

The authors have declared no competing interest.

### Funding Statement

This study was funded by the Villanova University M. Louise Fitzpatrick College of Nursing Research Development Grant.

### Author Declarations

IRB of the University of Pennsylvania gave ethical approval for this work

